# Machine Learning Estimation of Gestational Age at Delivery Using Linked Mother-Infant Electronic Health Records Across Two Health Systems

**DOI:** 10.64898/2026.05.23.26353959

**Authors:** Cosmin A. Bejan, Xiaotong Yang, Amelie Pham, Layth Qassem, Abin Abraham, Leena Choi, S. Trent Rosenbloom, Lana X. Gamire, Elizabeth J. Phillips

**Author notes:** **Corresponding authors**: Cosmin A. Bejan, PhD Department of Biomedical Informatics Vanderbilt University Medical Center 2525 West End Avenue, Suite 1500 Nashville, TN 37232, USA, Lana X. Garmire, PhD Department of Biomedical Informatics and Data Science University of Alabama at Birmingham ALGEN 428 701 19TH St South Birmingham, Alabama 35294, USA, Elizabeth J. Phillips, MD Department of Medicine Vanderbilt University Medical Center 1161 21st Ave S Nashville, TN 37232, USA.

## Abstract

**Objective:** This study aimed to train and evaluate supervised machine learning walgorithms using electronic health record (EHR) data to accurately estimate gestational age at delivery.

**Materials and Methods:** We trained random forest, gradient boosting, and ensemble models on EHR data of mother-infant dyads from Vanderbilt University Medical Center(VUMC) and replicated the analyses at University of Michigan (UMich). We further analyzed EHR predictors of gestational age, assessed temporal drift in EHR data elements, and evaluated model performance stratified by delivery status.

**Results:** The study included pregnancies corresponding to 54,344 and 34,345 mother-infant dyads at VUMC (2005-2025) and UMich (2012-2024), respectively. The gestational age predictions of the ensemble models achieved the highest agreement with the reference standard on the VUMC dataset (±1 week: 85.2%, ±2 weeks: 94.3%, MAE: 4.4 days) and demonstrated stronger generalization on the UMich dataset (±1 week: 93.1%, ±2 weeks: 97.8%, MAE: 2.8 days). Further, performance was better among pregnancies delivered in more recent years, and among full- and late-term deliveries compared with preterm deliveries.

**Discussion:** The results indicate that supervised machine learning methods leveraging linked mother-infant EHRs can accurately estimate gestational age at delivery, while demonstrating the generalizability of the modeling approach and the portability of the analytic workflow across healthcare sites.

**Conclusion:** This study presents a robust and generalizable machine learning framework to estimate gestational age at delivery. The framework can be reliably used to impute gestational age in large-scale, real-world clinical studies to support maternal and neonatal health research, in which accurate estimation of pregnancy onset is critical.

## 1. INTRODUCTION

Accurate estimation of gestational age is fundamental to pregnancy and offspring health outcome research. In pharmacoepidemiologic studies of drug safety in pregnancy, for example, accurate definition of pregnancy onset and exposure windows is particularly critical because fetal susceptibility to pharmacologic agents and teratogenic risk vary by developmental stage.^1–3^ However, errors in dating pregnancy onset are common and may occur for numerous reasons, including provider data entry errors, lack of ultrasound documentation, incomplete records when patients transfer care in pregnancy, or inadequate system infrastructure. Such inaccuracies in pregnancy dating can lead to misclassification of drug exposures, introducing bias and undermining the validity of study findings.^4–9^ These challenges are particularly pronounced in studies using electronic health record (EHR) and administrative claims databases. This is because observational studies based on these data sources are the primary method for evaluating the safety and effectiveness of treatments in pregnant populations who are typically excluded from randomized clinical trials due to design challenges and ethical concerns.^10–12^ In these data sources, gestational age estimates are often missing, incomplete, or incorrectly recorded. Such gaps may undermine statistical power in perinatal and pharmacoepidemiologic research involving pregnant populations, including drug-wide association studies,^13–15^ and may also introduce bias if missingness is related to confounding factors.

This study presents machine learning algorithms for estimating live birth gestational age at the time of delivery using EHR data of mother-infant dyads from two academic centers, Vanderbilt University Medical Center (VUMC) and the University of Michigan (UMich). While both centers routinely record clinical or obstetric gestational age estimates, approximately 9-10% of pregnancies from their EHR datasets have incomplete or invalid data for these estimates. The machine learning algorithms proposed in this work enable accurate estimation of gestational age in pregnancies with missing clinical estimates, offering a reliable solution for incorporating these pregnancies into analytic workflows. While prior research has primarily focused on estimating gestational age from administrative claims data,^16–19^ our methods demonstrate the added value of EHR-specific variables (e.g., infant birthweight and APGAR scores) and mother-infant linkage data for improving estimation accuracy. Additionally, our methods were developed and validated using EHR data derived from combined maternal-infant records and from maternal records alone in order to evaluate real-world scenarios corresponding to data sources with and without available mother-infant linkages, respectively.

## 2. METHODS

This is a retrospective cohort study that utilizes data pertaining to mother-infant dyads from the EHR systems of Vanderbilt University Medical Center (VUMC) and the University of Michigan (UMich) Medicine Healthcare System. The study involves the development and validation of machine learning algorithms to estimate gestational age at live birth for each pregnancy that has mother-infant dyad data. All machine learning models were initially trained on the VUMC dataset and subsequently retrained using the same modeling pipeline on the UMich dataset to assess the robustness and replicability of the development process. The study was approved with waiver of consent by the Institutional Review Board (IRB) at VUMC (IRB# 231322) and UMich (HUM# 00168171).

### 2.1 Data sources

Clinical data at VUMC were extracted from the Research Derivative (RD), a research-focused repository of the institution’s EHR system. The RD contains records on over 5.5 million patients, with mother-infant linkages available since 2005. To replicate the study, we used EHR data from the UMich database, which includes clinical records for approximately 5 million patients dating back to 2006. Specific data elements extracted from both EHR systems include demographics data, encounter information, clinical estimates of gestational age, APGAR scores, birthweight, and disease diagnoses based on the International Classification of Diseases, 9th/10th Revision, Clinical Modification (ICD-9/10-CM) billing codes.

### 2.2 Study population

The initial cohort included all mother-infant dyads from the EHR databases. To address within-mother correlation and potential bias, one infant per mother was randomly selected for mothers with multiple pregnancies or multiple births, resulting in approximately independent mother-infant dyads for model training. Further, to reduce potential sources of bias in the data, mothers who had inadequate prenatal care or pregnancies that did not result in a live birth were also excluded. Adequate prenatal care was defined using ICD diagnostic codes (**Table S1**) and the requirement of at least two prenatal encounters. Live births were identified using ICD codes documented at the time of delivery (**Table S2**). In summary, the final cohort used for algorithm development and validation consisted of unique mother-infant dyads with pregnancies resulting in live birth, in which mothers had adequate prenatal care and a documented clinical estimate of gestational age at delivery, as described below.

### 2.3 EHR-based extraction of gestational age estimates

Clinical gestational age (CGA) estimate at delivery was extracted from each maternal record and used as the reference standard outcome for training and validating the machine learning models. At VUMC, CGA estimates are recorded as textual patterns indicating weeks + days of gestation (e.g., “39 weeks 2/7” corresponds to 275 gestational days; more examples are provided in **Table S3**). All these textual expressions were transformed into a numeric representation of gestational age in days using regular expressions. Cases with no CGA expression within one day of delivery, or with expressions that could not be parsed (see examples in **Table S4**), were marked as missing or invalid, respectively.

### 2.4 Algorithm-based estimation of gestational age

We trained supervised machine learning models on mother-infant dyad data to predict the gestational age as a continuous numerical outcome. Specifically, to solve this regression task, we used random forest (RF), gradient boosting (GB), and an ensemble approach that involved averaging the predictions from the former two algorithms. First, we randomly split the mother-infant dyads into training (80%) and test (20%) sets. We then trained machine learning models and optimized hyperparameters using 5-fold cross-validation on the training set. **Table S5** summarizes the hyperparameters examined, the search ranges used, and the optimal values identified. We used a quantile transformation on the CGA values to address their left-skewed distribution and reduce the influence of outliers, which enhanced the performance of our regression models. Finally, we retrained the models on the full training set with the best-performing hyperparameter values identified through grid search and evaluated the retrained models on the test set. To provide a point of comparison for the machine learning models, we also implemented simple constant-value baselines using the mean and median CGA values from the training set.

For each mother-infant dyad, predictor variables used for model training were derived from three data sources: (1) pregnancy-related data from maternal records, (2) standard data elements commonly stored in EHR systems, and (3) pregnancy-related data from infant records. The EHR specific predictors included the infant birthweight, the 1-minute and 5-minutes APGAR scores, the number of previous pregnancies, and the number of fetuses in the current pregnancy associated with the mother-infant dyad. We addressed missing APGAR scores and infant birthweight values by imputing them with the respective median values. If multiple APGAR or birthweight values were recorded for the same infant, we selected the maximum value for each predictor. For the predictors extracted from mother records, we used ICD codes indicating preterm birth (**Table S6**), term or postterm birth (**Table S7**), fetal growth restriction (**Table S8**), and excessive fetal growth (**Table S9**). Similarly, from infant records, we extracted ICD-based predictors denoting preterm birth (**Table S10**), term or postterm birth (**Table S11**), small for gestational age (**Table S12**), and large for gestational age (**Table S13**). We also extracted predictors related to the demographic characteristics of the mother-infant dyad such as maternal age at delivery, maternal race and ethnicity, infant sex, and infant race and ethnicity. Following a pilot study on the training set, where ICD-based predictors were encoded as either binary or count variables at the individual code or broader category level, we determined that representing ICD codes by the number of distinct days or visits on which they were assigned during pregnancy was the most effective approach. To extract the ICD-based predictors for each mother-infant dyad, we estimated the pregnancy duration based on the delivery date (corresponding to the infant’s date of birth) and the median CGA value from the training set. Use of the dyad-specific CGA was avoided to prevent target leakage. Lastly, to evaluate the contribution of different predictor categories, we trained models using three configurations: (C1: Maternal), incorporating only predictors derived from maternal records; (C2: Maternal+EHR), adding EHR-derived features; and (C3: Maternal+EHR+Pediatric), including all available predictors. C1 and C2 correspond to data environments ranging from administrative databases to EHR systems without mother-infant linkage, while C3 represents environments with linked mother-infant data, which are more difficult to obtain in practice. **Table S14** summarizes the predictors used for each configuration.

### 2.5 Evaluation

We assessed algorithm performance over the test sets using the proportion of pregnancies with algorithm-based gestational age (AGA) estimation within ±1 week (±7 days) and ±2 weeks (±14 days) of the reference standard, and mean absolute error (MAE) between AGA and CGA estimates. To estimate the 95% confidence intervals (CIs) for these performance metrics, we applied bootstrapping by generating 1,000 resampled test sets with replacement, computing the metrics for each, and extracting the 2.5th and 97.5th percentiles of the resulting empirical distribution.^20^ Further, we investigated potential data drift arising from temporal shifts in ICD coding and billing practices by stratifying the results for pregnancies with delivery dates occurring before or on/after October 1, 2015, the date when the transition from ICD-9-CM to ICD-10-CM officially took place. Finally, we assessed variations in algorithm performance by delivery type, stratifying the results into preterm (less than 37 weeks of gestation), early term (37 0/7 weeks of gestation through 38 6/7 weeks of gestation), full term (39 0/7 weeks of gestation through 40 6/7 weeks of gestation), late term (41 0/7 weeks of gestation through 41 6/7 weeks of gestation), and postterm (42 0/7 weeks of gestation and beyond) categories.^21^

### 2.6 Method replication

To assess the robustness and reproducibility of our method development and determine whether comparable performance could be achieved, we independently replicated the machine learning framework for gestational age estimation on the UMich dataset. This replication used identical inclusion and exclusion criteria, predictor variables, and model training and evaluation procedures.

## 3. RESULTS

### 3.1 Selection of mother-infant dyads

The initial VUMC and UMich cohorts comprised 91,471 and 55,733 mother-infant dyads, corresponding to 68,659 and 39,969 mothers, respectively (**Figure 1**). The delivery years for dyads at VUMC and UMich spanned 2005-2025 and 2012-2024, respectively. During these periods, missing or invalid CGA values were most common in the early years following implementation of mother-infant linkages in both EHR systems (**Figures 2a-2b**). After retaining one infant per mother and excluding non-live births and mothers with insufficient prenatal care, the final analytic cohorts included 60,057 (65.7%) mother-infant dyads at VUMC and 37,861 (67.9%) at UMich. Dyads with valid CGA values in VUMC (N=54,344) and UMich (N=34,345) datasets were subsequently used for algorithm development (**Figure 1**). The average maternal age was 28.9 years in the VUMC dataset and 31.8 years in the UMich dataset (**Table 1**). Both datasets had predominantly White mothers and a slightly higher proportion of male infants. The CGA distributions were comparable, each with a median of 274 gestational days (**Table 1**, **Figures 2c-2d**).

**Figure 1.**
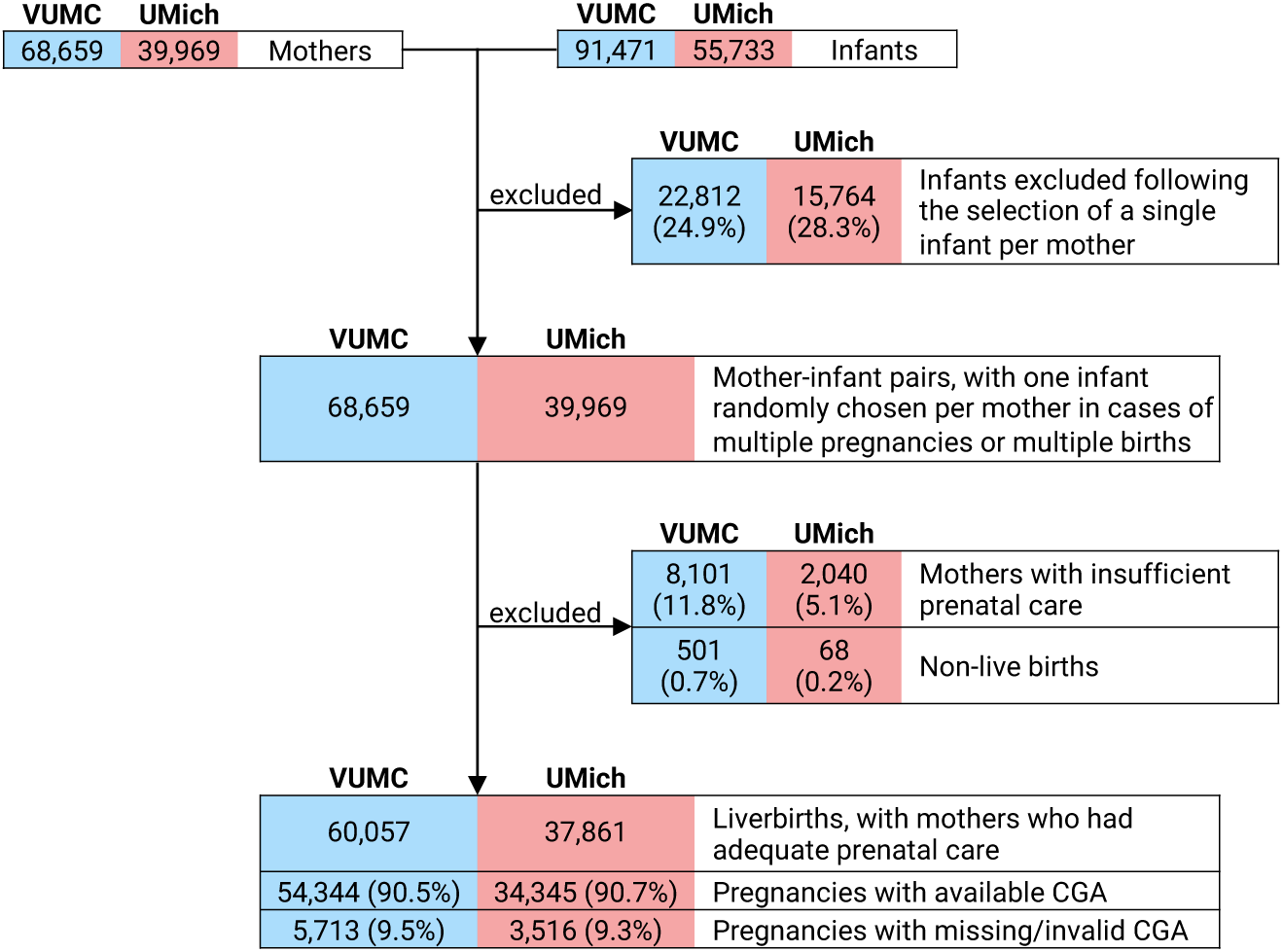
Selection of mother-infant dyads for algorithm-based estimation of gestational age in the electronic health record (EHR) systems of Vanderbilt University Medical Center (VUMC) and the University of Michigan (UMich) Medicine Healthcare System. CGA, clinical estimate of gestational age.

**Figure 2.**
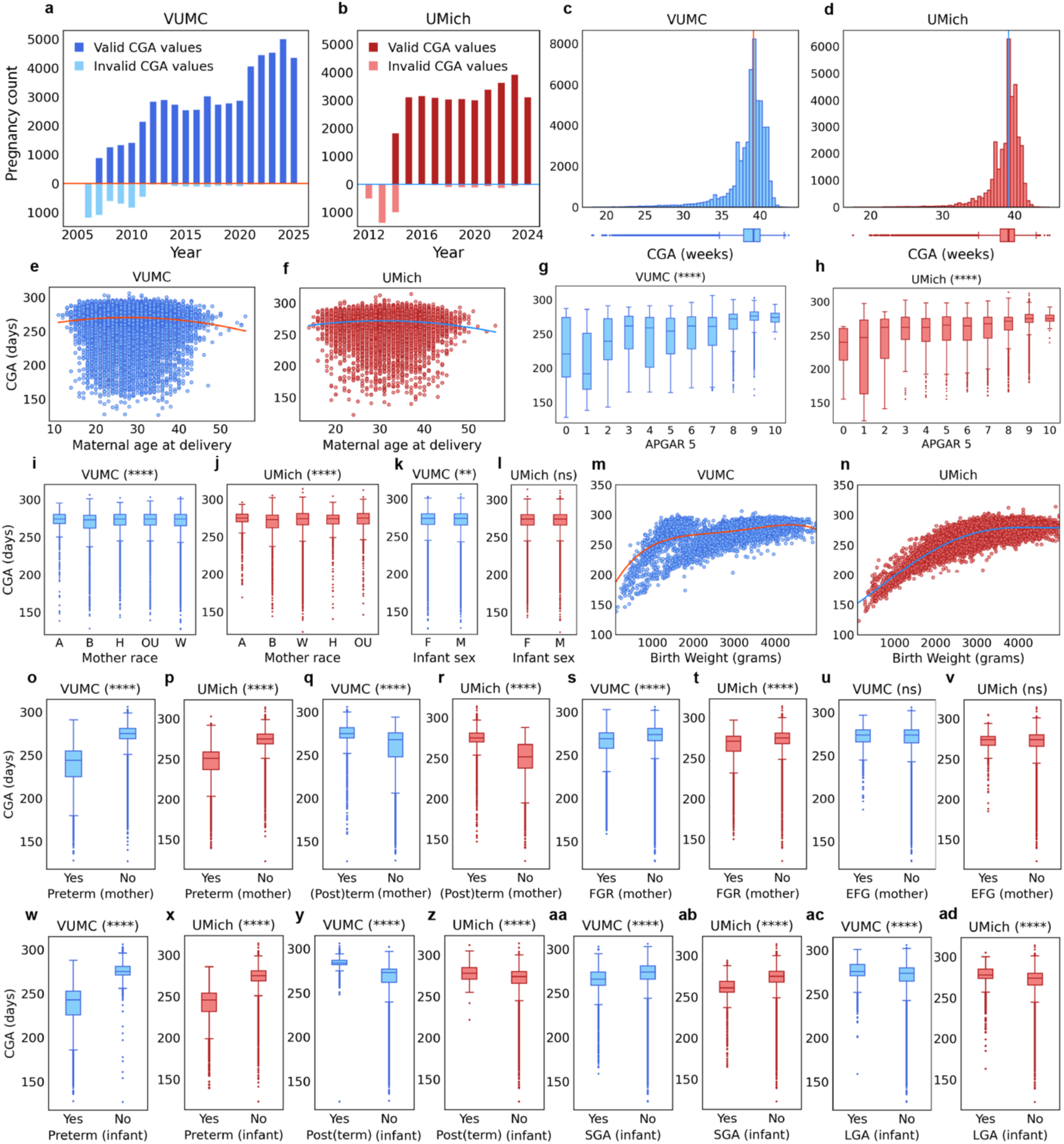
Comparative analysis of clinical gestational age (CGA) estimates across VUMC and UMich datasets. (**a-b**) Pregnancies with valid and invalid CGA estimates, by delivery year. (**c-d**) CGA distributions. The median CGA in both datasets is 274 days (39 1/7 weeks). (**e-f**) Maternal age at delivery and CGA. A scatterplot with quadratic fit was used to illustrate the nonlinear trend. (**g-h**) APGAR score at 5 minutes after delivery and CGA. (**i-j**) Maternal race and CGA. (**k-l**) Infant sex and CGA. (**m-n**) Birth weight and CGA. A smooth quartic spline was employed to visualize the trend. (**o-p**) Preterm in maternal records and CGA. (**q-r**) Term/postterm in maternal records and CGA. (**s-t**) Fetal growth restriction (FGR) in maternal records and CGA. (**u-v**) Excessive fetal growth (EFG) in maternal records and CGA. (**w-x**) Preterm in infant records and CGA. (**y-z**) Term/postterm in infant records and CGA. (**aa-ab**) Small for gestational age (SGA) in infant records and CGA. (**ac-ad**) Large for gestational age (LGA) in infant records and CGA. Predictor analysis (**e-ad**) was conducted using the dyads from the training set. Differences in CGA distributions were assessed using the Mann-Whitney U test for binary predictors and the Kruskal-Wallis H test for multicategorical predictors. Statistical significance levels are shown in the plot titles using the following notation: ‘****’ for *p*≤0.0001, ‘***’ for 0.0001<*p*≤0.001, ‘**’ for 0.001<*p*≤0.01, ‘*’ for 0.01<*p*≤0.05, and ‘ns’ for p>0.05.

**Table 1.** Characteristics of mother-infant dyads from VUMC and UMich datasets.

| Characteristic | VUMC | UMich |
| --- | --- | --- |
| Mother-infant dyads, N | 54,344 | 34,345 |
| Maternal age at delivery, mean years (SD) | 28.9 (5.8) | 31.8 (5.3) |
| **Mother race/ethnicity, N (%)** |  |  |
| White | 30,546 (56.2%) | 23,575 (68.6%) |
| Black | 7,648 (14.1%) | 4,146 (12.1%) |
| Hispanic | 8,387 (15.4%) | 1,984 (5.8%) |
| Asian | 2,101 (3.9%) | 3,130 (9.1%) |
| Other/unknown | 5,662 (10.4%) | 1,510 (4.4%) |
| **Infant sex, N (%)** |  |  |
| Male | 27,965 (51.5%) | 17,584 (51.2%) |
| Female | 26,379 (48.5%) | 16,761 (48.8%) |
| **CGA estimates in days** |  |  |
| Mean (SD) | 269.5 (18.8) | 270.9 (16.3) |
| Median | 274 | 274 |
| IQR | 265.0 - 280.0 | 266.0 - 280.0 |
| Min-Max | 127.0 - 306.0 | 123.0 - 314.0 |
| **Delivery status, N (%)** |  |  |
| Preterm | 7,795 (14.3%) | 4,336 (12.6%) |
| Early term | 14,090 (25.9%) | 8,874 (25.8%) |
| Full term | 27,881 (51.3%) | 18,333 (53.3%) |
| Late term | 4,350 (8.0%) | 2,644 (7.7%) |
| Postterm | 228 (0.4%) | 158 (0.5%) |

CGA, clinical gestational age; IQR, interquartile range; SD, standard deviation

### 3.2 Predictor analysis in association with CGA

A visual analysis highlighting differences in gestational age trends across selected predictors is shown in **Figures 2e-2ad**. The analysis was conducted using all mother-infant dyads from the VUMC (N= 43,475) and UMich (N=27,476) training sets. **Figures 2e-2f** indicate lower gestational age among young mothers and mothers of advanced maternal age in both cohorts. **Figures 2g-2h** demonstrate that higher APGAR scores at five minutes correspond to longer gestational ages. **Figures 2i-2j** show statistically significant variations in gestational age across maternal racial groups (*p*≤0.0001), with Asian mothers exhibiting the highest and Black mothers the lowest gestational ages at both sites (VUMC: 271.7 vs. 267.2 days; UMich: 273.0 vs. 268.1 days). Gestational age was slightly higher for mothers of female infants compared to male infants at VUMC (269.8 vs. 269.2 days; *p*<0.01), while no significant differences by infant sex were observed at UMich (**Figures 2k-2l**). Gestational age also showed a pronounced decrease within the first quartile of the birthweight distribution (**Figures 2m-2n**). As expected, dyads in which both maternal and infant records contain preterm ICD codes exhibited significantly lower gestational ages (**Figures 2o-2p; 2w-2x**), whereas those with term or postterm codes show significantly higher gestational ages (**Figures 2q-2r; 2y-2z**). In both datasets, gestational age was significantly lower for mothers with fetal growth restriction (FGR) ICD codes (VUMC: 262.3 vs. 270.6 days; UMich: 264.5 vs. 272.3 days; *p*≤0.0001) but did not differ significantly for those with excessive fetal growth (EFG) ICD codes (**Figures 2s-2v**). Finally, **Figures 2aa-2ad** indicate that gestational age was significantly lower for pregnancies with small for gestational age (SGA) ICD codes in infant records (VUMC: 262.7 vs. 270.0 days; UMich: 259.0 vs. 270.6; *p*≤0.0001) and higher for those with large for gestational age (LGA) ICD codes (VUMC: 275.3 vs. 269.1 days; UMich: 276.8 vs. 270.6 days; *p*≤0.0001).

### 3.3 CGA estimation performance

The results describing the performance of the machine learning algorithms for estimating gestational age on VUMC and UMich test sets are summarized in **Table 2** and **Figure 3**. When evaluated on the VUMC test set with the full set of predictors (C3), the algorithms correctly classified 84.0%-85.2% of pregnancies within 1 week and 93.7%-94.3% within 2 weeks of the reference standard. Retraining the algorithms on the UMich dataset using the same predictors yielded higher agreement with the reference standard, with 91.1%-93.1% of estimates within 1 week and 97.0%-97.8% within 2 weeks. Across both test sets, the machine learning algorithms substantially outperformed the baseline estimates based on the mean and median gestational ages from the training sets (VUMC, ±1 week: 41.5%-51.7%, ±2 weeks: 72.4%-80%; UMich, ±1 week: 42.7%-53%, ±2 weeks: 75.1%-81.6%). On the VUMC test set, the machine learning algorithms achieved MAE values ranging from 4.4 to 4.6 days, representing improvements of 6.8 to 7.9 days compared with the mean (12.3 days) and median (11.4 days) baselines. On the UMich test set, MAE values ranged from 2.8 to 3.1 days, corresponding to improvements of 7.3 to 8.2 days relative to these baselines (**Table 2**). Overall, the ensemble method achieved the best performance, followed in close succession by random forest and gradient boosting (**Table 2**, **Figures 3a-3b**). The predicted gestational ages from the three algorithms align closely with the reference estimates, as evidenced by substantial overlap in their distributions depicted in **Figures 3e-3f, 3i-3j, and 3m-3n** and the strong agreement shown in the scatterplots from **Figures 3g-3h, 3k-3l, and 3o-3p**, with Pearson correlation coefficients ranging from 0.86 to 0.89 for VUMC and 0.94 to 0.95 for UMich data. Across the three predictor configurations, model performance improves with each addition of data elements, from C1: maternal predictors alone to C2: maternal and EHR, and, finally, C3: maternal, EHR, and pediatric predictors (**Figures 3c-3d, Table S15**). The temporally stratified analysis indicates that overall model performance is improved for pregnancies delivered on or after October 1, 2015, compared with those delivered before (**Table S16**). Finally, the stratified analysis by delivery status shows that the models perform best for full-term and late-term deliveries, but substantially worse for preterm deliveries (**Table S17**).

**Figure 3.**
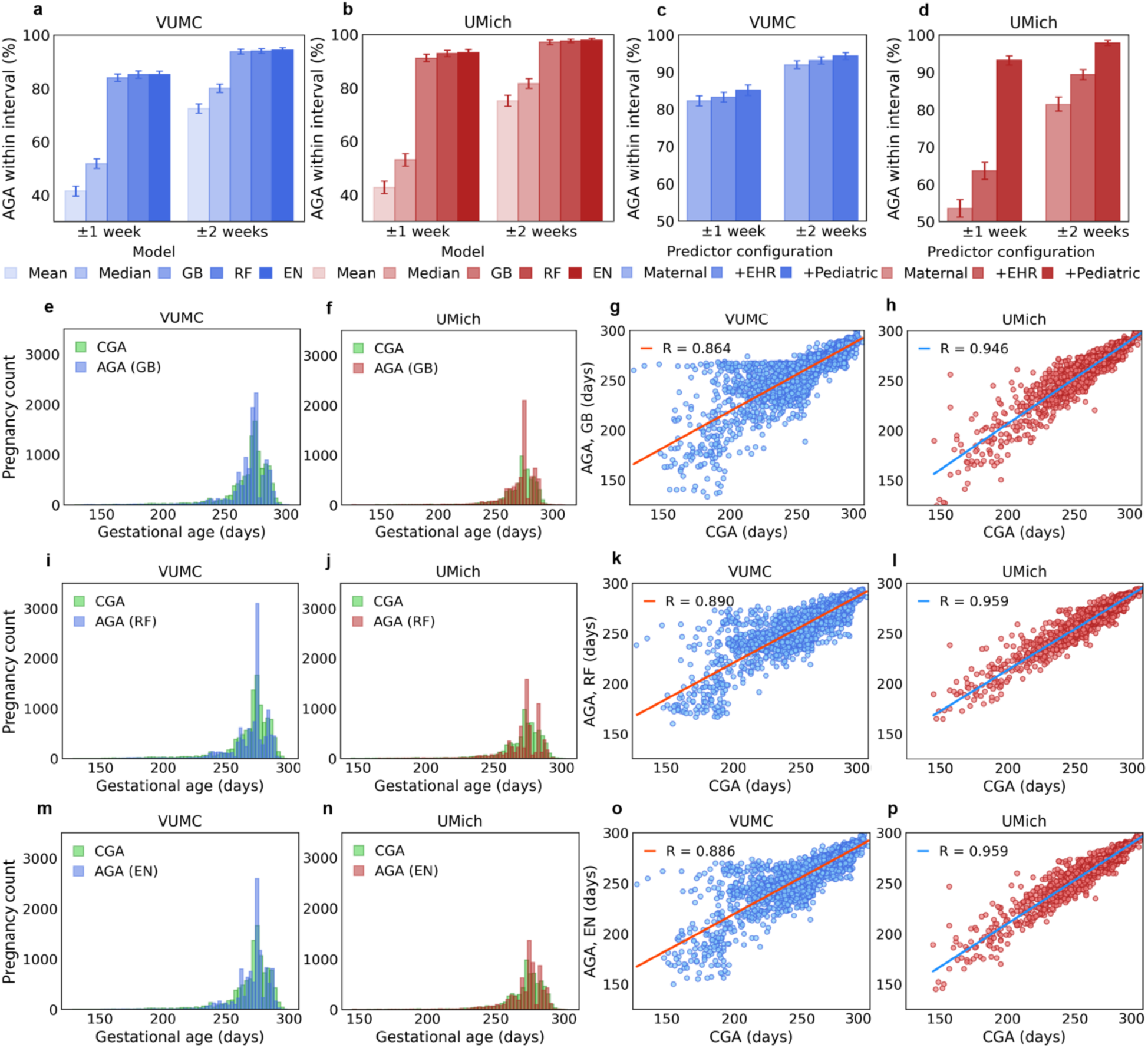
Performance evaluation of methods for gestational age prediction. (**a-b**) Comparative performance of baselines (mean and median) and machine learning methods measured by the proportion of gestational age predictions within 1 and 2 weeks of the reference standard. (**c-d**) Comparison of ensemble method performance using three predictor configurations: maternal only, maternal + EHR-specific, and maternal + pediatric + EHR-specific predictors. (**e-f**) Comparison of gestational age distributions between the reference standard and gradient boosting predictions. (**g-h**) Scatterplot of reference standard versus gradient boosting predictions. (**i-j**) Comparison of gestational age distributions between the reference standard and random forest predictions. (**k-l**) Scatterplot of reference standard versus random forest predictions. (**m-n**) Comparison of gestational age distributions between the reference standard and ensemble method predictions. (**o-p**) Scatterplot of reference standard versus ensemble method predictions. CGA, clinical gestational age; AGA, algorithm-derived gestational age; GB, gradient boosting; RF, random forest; EN, ensemble method; R, Pearson correlation coefficient.

**Table 2.** Algorithm performance evaluation on the VUMC and UMich test sets.

| Dataset | Dyads | Algorithm | % of AGA estimates<br>within 1 week (95% CI) | % of AGA estimates<br>within 2 weeks (95% CI) | MAE (95% CI) |
| --- | --- | --- | --- | --- | --- |
| VUMC | 10,869 | Mean | 41.5 (40.5-42.4) | 72.4 (71.5-73.3) | 12.3 (12.0-12.6) |
|  |  | Median | 51.7 (50.8-52.6) | 80.0 (79.3-80.8) | 11.4 (11.1-11.7) |
|  |  | Random Forest | 85.2 (84.5-85.9) | 93.9 (93.5-94.4) | 4.4 (4.2-4.5) |
|  |  | Gradient Boosting | 84.0 (83.3-84.6) | 93.7 (93.3-94.2) | 4.6 (4.5-4.8) |
|  |  | Ensemble | 85.1 (84.4-85.8) | 94.3 (93.9-94.8) | 4.4 (4.2-4.5) |
| UMich | 6,869 | Mean | 42.7 (41.6-43.9) | 75.1 (74.1-76.2) | 11.0 (10.7-11.3) |
|  |  | Median | 53.0 (51.8-54.2) | 81.6 (80.7-82.5) | 10.4 (10.1-10.7) |
|  |  | Random Forest | 92.9 (92.2-93.4) | 97.5 (97.1-97.8) | 2.8 (2.7-2.9) |
|  |  | Gradient Boosting | 91.1 (90.4-91.8) | 97.0 (96.6-97.4) | 3.1 (3.0-3.2) |
|  |  | Ensemble | 93.1 (92.5-93.7) | 97.8 (97.5-98.2) | 2.8 (2.7-2.9) |

AGA, algorithm-derived gestational age; MAE, mean absolute error; CI, confidence interval

## 4. DISCUSSION

Using EHR data from a cohort of mother-infant dyads, this study describes robust and accurate machine learning approaches for estimating gestational age at delivery. Applying the same machine learning framework, we independently reimplemented and retrained the models in a separate cohort, achieving improved performance. This successful cross-site replication demonstrates both the generalizability of the modeling approach and the portability of the analytic workflow.

Prior approaches to gestational age estimation primarily relied exclusively on administrative claims data, including methods that assume a uniform pregnancy duration and rule-based or machine learning models based largely on ICD code-derived predictors.^16–19,22–26^ Most were developed for live birth cohorts, with few addressing pregnancies ending in stillbirth, spontaneous abortion, or termination. The study most closely related to ours is by Zhu et al.,^24^ who used claims data from 114,117 pregnancies associated with mother-infant dyads in the Medicaid Analytic eXtract (MAX) to develop machine learning methods for estimating gestational age at live birth. Their best-performing model was an ensemble learning approach, Super Learner,^27^ which combined predictions from LASSO regression,^28^ Bayesian additive regression trees (BART),^29^ extreme gradient boosting,^30^ neural networks, Bayesian generalized linear models, generalized linear regression, and random forests, achieving 85.1% and 96.4% of estimates within 1 and 2 weeks of the reference standard, respectively. In comparison with prior administrative claims-based studies, including Zhu et al., our best-performing EHR-based approach demonstrated comparable or improved performance, achieving agreement within 1 and 2 weeks of 85.1% and 94.3% at VUMC and 93.1% and 97.8% at UMich.

This study exhibits several notable strengths. First, we conducted an analysis of gestational age patterns in relation to multiple EHR-derived predictors. This analysis revealed an inverted U-shaped relationship between gestational age and maternal age, consistent with prior literature describing similar^31,32^ or related U-shaped associations between maternal age and preterm birth risk.^33–36^ Additional findings corroborated by previous studies include associations between prematurity, defined as delivery before 37 weeks of gestation, low birthweight,^37,38^ small for gestational age,^39,40^ and fetal growth restriction.^41–43^ Second, we developed several methodological configurations that account for differences in predictor availability and mother-infant linkage, thereby supporting method implementation across diverse data environments, including administrative claims databases and EHR systems. Third, the temporally stratified analysis indicated that ICD-10-CM predictors contributed to substantially improved performance compared with ICD-9-CM predictors. In addition, stratification by delivery status showed that the machine learning algorithms performed optimally for full-term and late-term deliveries but were moderate for preterm deliveries, consistent with findings reported in prior studies.^16,17,19,24^ Finally, the independent reimplementation and retraining of the same machine learning framework at a second EHR site demonstrated the robustness and generalizability of the approach, as well as the portability of the analytic pipeline across institutions. The consistently high performance achieved across sites indicates that the learned associations are not specific to any single health system, patient population, or EHR implementation, but rather reflect broadly generalizable patterns. Notably, the performance was improved in the UMich cohort, potentially due to its reduced reliance on ICD-9-CM predictors. In contrast, the VUMC dataset includes a substantial number of pregnancies from earlier years (i.e., 2005-2011), which may be associated with greater gestational age measurement error due to more limited ultrasound technology (e.g., lower-resolution imaging) and less advanced modeling methodologies available at the time.

The system also exhibits several limitations that warrant further investigation and refinement in future research. Notably, the machine learning methods demonstrated suboptimal performance for preterm deliveries. Future research will explore incorporating additional predictors derived from maternal and infant records, including a broader range of ICD codes, Current Procedural Terminology (CPT) codes, and Neonatal Intensive Care Unit (NICU)-related data elements. Further, gestational age estimation for pregnancies ending in stillbirth, ectopic pregnancy, spontaneous abortion, or termination was not investigated, as nearly all pregnancies in both VUMC and UMich cohorts were associated with live birth ICD codes. However, further evaluation is needed to determine whether using the same set of predictors within the proposed machine learning framework will produce similar results on datasets in which these outcomes are available. Additionally, the potential for machine learning-based gestational age prediction to perpetuate or exacerbate health disparities remains unexplored. Although beyond the scope of this study, a systematic evaluation of model fairness, along with the application of bias mitigation techniques, will be essential to ensure transparency and equity. In this context, biased models may systematically misestimate gestational age for certain groups, leading to inaccurate pregnancy onset dating, exposure misclassification, and underrepresentation in pharmacoepidemiologic pregnancy research.

## 5. CONCLUSION

This study shows that machine learning models trained on EHR data can accurately and robustly estimate gestational age at delivery. The novelty of this work lies in leveraging EHR-derived predictors and mother-infant linkage information within a supervised machine learning framework, demonstrating both the generalizability of the modeling approach and the portability of the analytic workflow across EHR sites. These findings indicate that the models can be reliably used as imputation tools for large-scale real-world clinical studies involving pregnant populations across heterogeneous data sources, including administrative claims databases and EHR systems with partially or completely missing gestational age information. Continued efforts are needed to improve model performance in estimating gestational age for preterm deliveries and to address and mitigate potential biases.

## Data availability

The code used in the analysis is available on GitHub at https://github.com/bejanlab/ML4EGA.git. The summary statistics extracted for this study are provided in the manuscript and supplementary material. Any request to access the Research Derivative data will need to be reviewed and approved by Vanderbilt University Medical Center. Researchers will need to provide evidence of IRB approval for their study. For the approved studies, data will be released via a Data Use Agreement. Interested investigators who meet the criteria for accessing sensitive data can contact the University of Michigan Precision Health’s Research Scientific Facilitators at (also see https://research.medicine.umich.edu/our-units/data-office-clinical-translational-research/data-access) to inquire about the UMich dataset and the necessary steps regarding ethics committee approval and data accessing agreement.

## Funding

CAB, AP, LC, TR, and EJP are supported by R21HD113234. XY is supported by T32GM141746. LXG is supported by NIH grants R01LM012373, R01LM012907, R03OD039978 and R01HD084633.

## Author Contributions

CAB initiated the project, developed the code, and analyzed the VUMC data. XY replicated the study and analyzed the UMich data. LXG provided the UMich data and supervised XY on the replication study. CAB and XY had full responsibility for the integrity of the data and the accuracy of the data analysis.

*Concept and design:* CAB, AP, LC, STR, EJP.

*Acquisition, analysis, or interpretation of data:* CAB, XY, AP, LQ, AA, LC, STR, LXG, EJP.

*Drafting of the manuscript:* CAB.

*Critical review of the manuscript for important intellectual content:* All authors.

*Statistical analysis:* CAB.

*Obtained funding:* CAB, LXG.

*Administrative, technical, or material support:* XY, AP, LQ, LC, STR, LXG, EJP.

*Supervision:* CAB, LXG, EJP.

## Disclosures

AP receives industry-sponsored research grant funding from the Novocuff medical device company. She also serves as a member of the Data and Safety Monitoring Board (DSMB) for an NIH-funded study at Vanderbilt University Medical Center (NIH Grant Number: 1R01MH136399). EJP has received consulting fees and royalties from UpToDate®, consulting fees from Janssen, AstraZeneca, Intellia, Verve and Glenmark and grant funding from the NIH (R01HG010863, R01AI152183, U01AI154659, R13AR085978), National Health and Medical Research Council of Australia (NHMRC) and Leo Foundation unrelated to the published work. No other conflicts of interest are reported.

## Notes

### Author Declarations

The institutional review boards at Vanderbilt University Medical Center and the University of Michigan Medicine Healthcare System gave ethical approval for this study.

