## Supplemental Tables for "Machine Learning Estimation of Gestational Age at Delivery Using Linked Mother-Infant Electronic Health Records Across Two Health Systems"

**Table S1** Insufficient prenatal care ICD codes from maternal records.

**Table S2** Live birth ICD codes from maternal and infant records.

**Table S3** Text expressions representing valid clinical estimates of gestational age in the EHR.

**Table S4** Unparseable text expressions for extracting accurate gestational age values in days.

**Table S5** Hyperparameters used for optimizing the Random Forest and Gradient Boosting models.

**Table S6** Preterm ICD codes from maternal records.

**Table S7** Term or postterm ICD codes from maternal records.

**Table S8** Fetal growth restriction (FGR) ICD codes from maternal records.

**Table S9** Excessive fetal growth (EFG) ICD codes from maternal records.

**Table S10** Preterm ICD codes from infant records.

**Table S11** Term or postterm ICD codes from infant records.

**Table S12** Small for gestational age (SGA) ICD codes from infant records.

**Table S13** Large for gestational age (LGA) ICD codes from infant records.

**Table S14** Overview of predictors for each mother-infant dyad, categorized by data source.

**Table S15** Algorithm performance results using predictors from maternal records alone or in combination with EHR-specific data elements.

**Table S16** Performance results from the temporal stratification of pregnancies delivered before vs. on or after October 1, 2015.

**Table S17** Evaluation of algorithm performance stratified by delivery status.

**Table S1** Insufficient prenatal care ICD codes from maternal records.

| Code | Description |
|---|---|
| **ICD-9-CM** | |
| V23.7 | Supervision of high-risk pregnancy with insufficient prenatal care |
| **ICD-10-CM** | |
| O09.3xx | Supervision of pregnancy with insufficient antenatal care |

**Table S2** Live birth ICD codes from maternal and infant records.

| Code | Description |
|---|---|
| **ICD-9-CM** | |
| 765.1x | Other preterm infants |
| 766.0 | Exceptionally large baby |
| 766.1 | Other "heavy-for-dates" infants |
| 766.2x | Late infant |
| V27.0 | Outcome of delivery, single liveborn |
| V27.2 | Outcome of delivery, twins, both liveborn |
| V27.5 | Outcome of delivery, other multiple birth, all liveborn |
| V30.xx | Single liveborn |
| V31.xx | Twin, mate liveborn |
| V34.xx | Other multiple, mates all liveborn |
| V39.xx | Liveborn, unspecified whether single, twin or multiple |
| **ICD-10-CM** | |
| P07.0x | Extremely low birth weight newborn |
| P07.1x | Other low birth weight newborn |
| P07.3x | Preterm [premature] newborn |
| P08.0 | Exceptionally large newborn baby |
| P08.1 | Other heavy for gestational age newborn |
| P08.2 | Late newborn, not heavy for gestational age |
| P08.21 | Post-term newborn |
| P08.22 | Prolonged gestation of newborn |
| Z37.0 | Single live birth |
| Z37.2 | Twins, both liveborn |
| Z37.5x | Other multiple births, all liveborn |
| Z38.xx | Single or multiple liveborn infants according to place of birth and type of delivery |

**Table S3** Text expressions representing valid clinical estimates of gestational age in the EHR.

| | | | | |
|---|---|---|---|---|
| 40 weeks | 36.5 | 28 weeks 3 days | 38 weeks 2/7 | 41wks |
| 40 4/7 weeks | 36w2d | 37/5 | 36.5 weeks EGA | 40 wks. |
| 38w 2d | 35 5/7 days | 41 3/7wks | 38 2/7 weeks2 | 42weeks |
| 40 4/7 | 37 4/7weeks | 37.4 wks | 29 + 6weeks | 40weeks + 2 days |
| 40 | 37.4weeks | 34 weeks 0/7 days | 32 and 4/7 weeks | 40 weeks 1 day |
| 39w | 37weeks | 39 wks | 41 weeks & 1 day | 40 weeks and 2 days |
| 44 4/7 wks | 37+2 | 27weeks 1day | 39wks 5days | 41/1weeks |
| 41.1 weeks | 39/1 weeks | 40+1 weeks | 41 and 4 weeks | 39 1 /7 weeks |

**Table S4** Unparseable text expressions for extracting accurate gestational age values in days.

| | | | |
|---|---|---|---|
| 36-39 | Delivered | ~ 35 | ~42 weeks |
| 33-35 wks | no dating | 10/7/09 | 41 60/7 weeks |
| PP | unknown, term | unknown, est 34-36 by pt | approx 32 weeks |

**Table S5** Hyperparameters used for optimizing the random forest and gradient boosting models.

| Model hyperparameter | Value range | Optimal values on VUMC dataset | Optimal values on UMich dataset |
|---|---|---|---|
| **Random Forest** | | | |
| n_estimators | [100, 200] | 200 | 200 |
| max_depth | [None, 10, 20] | None | None |
| min_samples_split | [2, 5] | 5 | 2 |
| min_samples_leaf | [1, 2] | 1 | 1 |
| max_features | ['sqrt', 'log2'] | sqrt | sqrt |
| **Gradient Boosting** | | | |
| n_estimators | [100, 200] | 200 | 200 |
| learning_rate | [0.01, 0.1, 0.2] | 0.2 | 0.2 |
| max_depth | [3, 5, 7] | 7 | 7 |
| min_samples_split | [2, 5] | 5 | 2 |
| min_samples_leaf | [1, 3] | 1 | 3 |
| subsample | [0.8, 1.0] | 1.0 | 1.0 |
| max_features | ['sqrt', 'log2'] | sqrt | sqrt |

**Table S6** Preterm ICD codes from maternal records.

| Code | Description |
|---|---|
| **ICD-9-CM** | |
| 644.2x | Early onset of delivery |
| **ICD-10-CM** | |
| O42.01x | Preterm premature rupture of membranes, onset of labor within 24 hours of rupture |
| O42.11x | Preterm premature rupture of membranes, onset of labor more than 24 hours following rupture |
| O42.91x | Preterm premature rupture of membranes, unspecified as to length of time between rupture and onset of labor |
| O60.1 | Preterm labor with preterm delivery |
| O60.10xx | Preterm labor with preterm delivery, unspecified trimester |
| O60.12xx | Preterm labor second trimester with preterm delivery second trimester |
| O60.13xx | Preterm labor second trimester with preterm delivery third trimester |
| O60.14xx | Preterm labor third trimester with preterm delivery third trimester |

**Table S7** Term or postterm ICD codes from maternal records.

| Code | Description |
|---|---|
| **ICD-9-CM** | |
| 645 | Late pregnancy |
| 645.0x | Prolonged pregnancy |
| 645.1x | Post term pregnancy |
| 645.2x | Prolonged pregnancy |
| **ICD-10-CM** | |
| O42.92 | Full-term premature rupture of membranes, unspecified as to length of time between rupture and onset of labor |
| O48 | Late pregnancy |
| O48.0 | Post-term pregnancy |
| O48.1 | Prolonged pregnancy |
| O60.20Xx | Term delivery with preterm labor, unspecified trimester |
| O60.22Xx | Term delivery with preterm labor, second trimester |
| O60.23Xx | Term delivery with preterm labor, third trimester |
| O80 | Encounter for full-term uncomplicated delivery |
| Z3A.37 | 37 weeks gestation of pregnancy |
| Z3A.38 | 38 weeks gestation of pregnancy |
| Z3A.39 | 39 weeks gestation of pregnancy |
| Z3A.4 | Weeks of gestation of pregnancy, weeks 40 or greater |
| Z3A.40 | 40 weeks gestation of pregnancy |
| Z3A.41 | 41 weeks gestation of pregnancy |
| Z3A.42 | 42 weeks gestation of pregnancy |
| Z3A.49 | Greater than 42 weeks gestation of pregnancy |

**Table S8** Fetal growth restriction (FGR) ICD codes from maternal records.

| Code | Description |
| --- | --- |
| **ICD-9-CM** | |
| 656.5x | Poor fetal growth affecting management of mother |
| V28.4 | Antenatal screening for fetal growth retardation using ultrasonics |
| **ICD-10-CM** | |
| O36.5 | Maternal care for known or suspected poor fetal growth |
| O36.51 | Maternal care for known or suspected placental insufficiency |
| O36.511x | Maternal care for known or suspected placental insufficiency, first trimester |
| O36.512x | Maternal care for known or suspected placental insufficiency, second trimester |
| O36.513x | Maternal care for known or suspected placental insufficiency, third trimester |
| O36.519x | Maternal care for known or suspected placental insufficiency, unspecified trimester |
| O36.59 | Maternal care for other known or suspected poor fetal growth |
| O36.591x | Maternal care for other known or suspected poor fetal growth, first trimester |
| O36.592x | Maternal care for other known or suspected poor fetal growth, second trimester |
| O36.593x | Maternal care for other known or suspected poor fetal growth, third trimester |
| O36.599x | Maternal care for other known or suspected poor fetal growth, unspecified trimester |
| Z36.4 | Encounter for antenatal screening for fetal growth retardation |

**Table S9** Excessive fetal growth (EFG) ICD codes from maternal records.

| Code | Description |
| --- | --- |
| **ICD-9-CM** | |
| 656.6x | Excessive fetal growth affecting management of mother |
| **ICD-10-CM** | |
| O36.60xx | Maternal care for excessive fetal growth, unspecified trimester |
| O36.61xx | Maternal care for excessive fetal growth, first trimester |
| O36.62xx | Maternal care for excessive fetal growth, second trimester |
| O36.63xx | Maternal care for excessive fetal growth, third trimester |
| Z36.88 | Encounter for antenatal screening for fetal macrosomia |

**Table S10** Preterm ICD codes from infant records.

| Code | Description |
|---|---|
| **ICD-9-CM** | |
| 765 | Disorders relating to short gestation and unspecified low birthweight |
| 765.0x | Extreme immaturity |
| 765.1x | Other preterm infants |
| 774.2 | Neonatal jaundice associated with preterm delivery |
| **ICD-10-CM** | |
| P07 | Disorders of newborn related to short gestation and low birth weight, not elsewhere classified |
| P07.0x | Extremely low birth weight newborn |
| P07.1x | Other low birth weight newborn |
| P07.2x | Extreme immaturity of newborn |
| P07.3x | Preterm [premature] newborn [other] |
| P59.0 | Neonatal jaundice associated with preterm delivery |

**Table S11** Term or postterm ICD codes from infant records.

| Code | Description |
|---|---|
| **ICD-9-CM** | |
| 765.29 | 37 or more completed weeks of gestation |
| 766.2 | Late infant, not "heavy-for-dates" |
| 766.21 | Post-term infant |
| 766.22 | Prolonged gestation of infant |
| **ICD-10-CM** | |
| P08.2 | Late newborn, not heavy for gestational age |
| P08.21 | Post-term newborn |
| P08.22 | Prolonged gestation of newborn |

**Table S12** Small for gestational age (SGA) ICD codes from infant records.

| Code | Description |
|---|---|
| **ICD-9-CM** | |
| 764 | Slow fetal growth and fetal malnutrition |
| 764.0x | ‘Light-for-dates’ without mention of fetal malnutrition |
| 764.1x | ‘Light-for-dates’ with signs of fetal malnutrition |
| 764.2x | Fetal malnutrition without mention of ‘light-for-dates’ |
| 764.9x | Fetal growth retardation, unspecified |
| **ICD-10-CM** | |
| P05 | Disorders of newborn related to slow fetal growth and fetal malnutrition |
| P05.0x | Newborn light for gestational age |
| P05.1x | Newborn small for gestational age |
| P05.2 | Newborn affected by fetal (intrauterine) malnutrition not light or small for gestational age |
| P05.9 | Newborn affected by slow intrauterine growth, unspecified |

**Table S13** Large for gestational age (LGA) ICD codes from infant records.

| Code | Description |
|---|---|
| **ICD-9-CM** | |
| 766 | Disorders relating to long gestation and high birthweight |
| 766.0 | Exceptionally large baby |
| 766.1 | Other "heavy-for-dates" infants |
| **ICD-10-CM** | |
| P08 | Disorders of newborn related to long gestation and high birth weight |
| P08.0 | Exceptionally large newborn baby |
| P08.1 | Other heavy for gestational age newborn |

**Table S14** Overview of predictors for each mother-infant dyad, categorized by data source.

| Predictor category | Description |
|---|---|
| **Maternal** | Maternal age at delivery |
| | Maternal race and ethnicity |
| | Preterm ICD codes from maternal records |
| | Term or postterm ICD codes from maternal records |
| | Fetal growth restriction ICD codes from maternal records |
| | Excessive fetal growth ICD codes from maternal records |
| **EHR** | Number of previous pregnancies |
| | Number of fetuses in the current pregnancy |
| | The APGAR score at 1 minute after birth from maternal records |
| | The APGAR score at 5 minutes after birth from maternal records |
| | Infant weight at birth from maternal records |
| **Pediatric** | Infant sex |
| | Infant race and ethnicity |
| | Preterm ICD codes from infant records |
| | Term or postterm ICD codes from infant records |
| | Small for gestational age ICD codes from infant records |
| | Large for gestational age ICD codes from infant records |

**Table S15** Algorithm performance results using predictors from maternal records alone or in combination with EHR-specific data elements.

| Dataset | Dyads | Predictor category | Algorithm | % within 1 week (95% CI) | % within 2 weeks (95% CI) | MAE (95% CI) |
|---|---|---|---|---|---|---|
| VUMC | 10,869 | Maternal | Random Forest | 82.1 (81.4-82.8) | 91.6 (91.1-92.1) | 5.2 (5.0-5.4) |
| | | | Gradient Boosting | 81.1 (80.3-81.8) | 91.4 (90.8-91.9) | 5.5 (5.3-5.7) |
| | | | Ensemble | 82.3 (81.6-83.0) | 91.9 (91.4-92.4) | 5.2 (5.0-5.4) |
| | | Maternal + EHR | Random Forest | 83.1 (82.4-83.8) | 92.6 (92.1-93.1) | 4.7 (4.6-4.9) |
| | | | Gradient Boosting | 82.3 (81.6-83.1) | 92.5 (92.1-93.0) | 4.9 (4.8-5.1) |
| | | | Ensemble | 83.2 (82.5-83.9) | 93.1 (92.6-93.6) | 4.7 (4.6-4.9) |
| UMich | 6,869 | Maternal | Random Forest | 88.1 (87.3-88.8) | 94.1 (93.5-94.6) | 4.1 (3.9-4.3) |
| | | | Gradient Boosting | 86.9 (86.1-87.6) | 93.8 (93.2-94.3) | 4.2 (4.1-4.4) |
| | | | Ensemble | 87.8 (87.0-88.5) | 94.2 (93.7-94.7) | 4.0 (3.9-4.2) |
| | | Maternal + EHR | Random Forest | 90.6 (89.9-91.2) | 96.2 (95.8-96.7) | 3.2 (3.1-3.3) |
| | | | Gradient Boosting | 89.7 (89.0-90.4) | 96.4 (96.0-96.8) | 3.2 (3.1-3.3) |
| | | | Ensemble | 90.9 (90.2-91.5) | 96.8 (96.4-97.2) | 3.1 (3.0-3.2) |

MAE, mean absolute error; CI, confidence interval

**Table S16** Performance results from the temporal stratification of pregnancies delivered before vs. on or after October 1, 2015.

| Dataset | Delivery date | Dyads | Algorithm | % within 1 week (95% CI) | % within 2 weeks (95% CI) | MAE (95% CI) |
|---|---|---|---|---|---|---|
| VUMC | Before 10/01/2015 | 3,454 | Random Forest | 77.6 (76.3-79.0) | 95.3 (94.6-96.0) | 5.1 (4.9-5.3) |
| | | (31.8%) | Gradient Boosting | 76.5 (75.1-78.0) | 96.2 (95.5-96.8) | 5.0 (4.9-5.2) |
| | | | Ensemble | 77.6 (76.2-79.1) | 96.2 (95.6-96.8) | 4.9 (4.7-5.1) |
| | On/after 10/01/2015 | 7,415 | Random Forest | 88.7 (88.0-89.3) | 93.3 (92.7-93.9) | 4.0 (3.8-4.2) |
| | | (68.2%) | Gradient Boosting | 87.4 (86.7-88.2) | 92.6 (92.0-93.1) | 4.5 (4.2-4.7) |
| | | | Ensemble | 88.6 (87.9-89.3) | 93.4 (92.8-93.9) | 4.1 (3.9-4.3) |
| UMich | Before 10/01/2015 | 828 | Random Forest | 79.6 (76.7-82.4) | 96.9 (95.5-97.9) | 4.7 (4.4-5.1) |
| | | (12.1%) | Gradient Boosting | 79.9 (77.3-82.5) | 97.1 (95.9-98.2) | 4.7 (4.4-5.0) |
| | | | Ensemble | 80.9 (78.3-83.5) | 97.4 (96.4-98.6) | 4.6 (4.3-4.9) |
| | On/after 10/01/2015 | 6,041 | Random Forest | 94.7 (94.1-95.2) | 97.6 (97.2-98.0) | 2.6 (2.5-2.7) |
| | | (87.9%) | Gradient Boosting | 92.7 (92.0-93.3) | 97.0 (96.6-97.5) | 2.9 (2.8-3.0) |
| | | | Ensemble | 94.8 (94.2-95.4) | 97.9 (97.5-98.2) | 2.5 (2.4-2.6) |

MAE, mean absolute error; CI, confidence interval

**Table S17** Evaluation of algorithm performance stratified by delivery status.

| Dataset | Delivery status | Dyads N (%) | Algorithm | % within 1 week (95% CI) | % within 2 weeks (95% CI) | MAE (95% CI) |
|---|---|---|---|---|---|---|
| VUMC | Preterm | 1,569 | Random Forest | 39.5 (36.9-41.9) | 64.9 (62.5-67.4) | 14.5 (13.7-15.2) |
| | | (14.4%) | Gradient Boosting | 33.9 (31.5-36.4) | 60.3 (58.0-62.7) | 16.2 (15.4-17.0) |
| | | | Ensemble | 39.8 (37.3-42.1) | 65.1 (62.8-67.5) | 14.6 (13.9-15.4) |
| | Early term | 2,839 | Random Forest | 87.6 (86.3-88.7) | 98.7 (98.2-99.1) | 3.4 (3.2-3.5) |
| | | (26.1%) | Gradient Boosting | 88.5 (87.3-89.6) | 99.2 (98.8-99.5) | 3.4 (3.3-3.5) |
| | | | Ensemble | 88.3 (87.1-89.4) | 99.1 (98.8-99.5) | 3.3 (3.2-3.4) |
| | Full term | 5,551 | Random Forest | 94.8 (94.2-95.3) | 98.8 (98.5-99.1) | 2.4 (2.4-2.5) |
| | | (51.1%) | Gradient Boosting | 93.8 (93.2-94.5) | 99.4 (99.2-99.6) | 2.5 (2.4-2.5) |
| | | | Ensemble | 94.3 (93.7-94.9) | 99.2 (99.0-99.4) | 2.4 (2.3-2.5) |
| | Late term | 866 | Random Forest | 98.1 (97.2-99.0) | 99.7 (99.2-100.0) | 1.7 (1.6-1.8) |
| | | (8.0%) | Gradient Boosting | 96.6 (95.4-97.8) | 99.4 (98.8-99.9) | 1.8 (1.6-1.9) |
| | | | Ensemble | 97.8 (96.8-98.7) | 99.5 (99.1-99.9) | 1.7 (1.6-1.9) |
| | Postterm | 44 | Random Forest | 86.2 (75.0-95.5) | 100.0 (100.0-100.0) | 4.9 (4.2-5.7) |
| | | (0.4%) | Gradient Boosting | 81.7 (70.5-93.2) | 97.6 (93.2-100.0) | 4.8 (3.8-5.8) |
| | | | Ensemble | 83.8 (72.7-93.2) | 100.0 (100.0-100.0) | 4.5 (3.6-5.4) |
| UMich | Preterm | 901 | Random Forest | 64.2 (61.0-67.4) | 83.0 (80.6-85.6) | 7.8 (7.2-8.4) |
| | | (13.1%) | Gradient Boosting | 54.8 (51.6-58.0) | 79.0 (76.4-81.6) | 8.9 (8.4-9.5) |
| | | | Ensemble | 66.4 (63.4-69.4) | 84.5 (82.1-86.7) | 7.4 (6.9-8.0) |
| | Early term | 1,734 | Random Forest | 94.8 (93.8-95.8) | 99.4 (99.0-99.7) | 2.5 (2.4-2.6) |
| | | (25.2%) | Gradient Boosting | 93.7 (92.5-94.8) | 99.4 (99.0-99.8) | 2.9 (2.8-3.0) |
| | | | Ensemble | 95.0 (94.0-96.0) | 99.6 (99.3-99.9) | 2.6 (2.5-2.7) |
| | Full term | 3,636 | Random Forest | 98.1 (97.6-98.5) | 99.9 (99.7-100.0) | 1.9 (1.9-2.0) |
| | | (52.9%) | Gradient Boosting | 97.8 (97.3-98.3) | 99.9 (99.8-100.0) | 2.0 (1.9-2.0) |
| | | | Ensemble | 97.9 (97.4-98.4) | 99.9 (99.8-100.0) | 1.9 (1.9-2.0) |
| | Late term | 572 | Random Forest | 99.5 (98.8-100.0) | 100.0 (100.0-100.0) | 1.4 (1.3-1.6) |
| | | (8.3%) | Gradient Boosting | 98.9 (98.1-99.7) | 100.0 (100.0-100.0) | 1.4 (1.3-1.6) |
| | | | Ensemble | 99.5 (98.8-100.0) | 100.0 (100.0-100.0) | 1.4 (1.3-1.5) |
| | Postterm | 26 | Random Forest | 92.3 (80.8-100.0) | 100.0 (100.0-100.0) | 3.8 (2.9-4.8) |
| | | (0.4%) | Gradient Boosting | 80.9 (65.4-96.2) | 96.2 (88.5-100.0) | 3.6 (2.3-5.1) |
| | | | Ensemble | 92.3 (80.8-100.0) | 100.0 (100.0-100.0) | 2.7 (1.7-3.9) |

MAE, mean absolute error; CI, confidence interval
